# Diminishing immune responses against variants of concern in dialysis patients four months after SARS-CoV-2 mRNA vaccination

**DOI:** 10.1101/2021.08.16.21262115

**Authors:** Alex Dulovic, Monika Strengert, Gema Morillas Ramos, Matthias Becker, Johanna Griesbaum, Daniel Junker, Karsten Lürken, Andrea Beigel, Eike Wrenger, Gerhard Lonnemann, Anne Cossmann, Metodi V. Stankov, Alexandra Dopfer-Jablonka, Philipp D. Kaiser, Bjoern Traenkle, Ulrich Rothbauer, Gérard Krause, Nicole Schneiderhan-Marra, Georg M.N. Behrens

**Affiliations:** NMI Natural and Medical Sciences Institute at the University of Tübingen, Reutlingen, Germany; Helmholtz Centre for Infection Research, Braunschweig, Germany; TWINCORE GmbH, Centre for Experimental and Clinical Infection Research, a joint venture of the Hannover Medical School and the Helmholtz Centre for Infection Research, Hannover, Germany; Department for Rheumatology and Immunology, Hannover Medical School, Hannover, Germany; Dialysis Centre Eickenhof, Langenhagen, Germany; German Centre for Infection Research (DZIF), partner site Hannover-Braunschweig, Germany; Pharmaceutical Biotechnology, University of Tübingen, Germany; CiiM - Centre for Individualized Infection Medicine, Hannover, Germany

## Abstract

Patients undergoing chronic hemodialysis were among the first to receive SARS-CoV-2 vaccinations due to their increased risk for severe COVID-19 disease and high case fatality rates. To date, there have been minimal longitudinal studies in hemodialysis patients to ascertain whether protection offered by vaccination is long-lasting. To assess how surrogates for protection changed over time, we examined both the humoral and cellular response in a previously reported cohort of at-risk hemodialysis patients and healthy donors, four months after their second dose of Pfizer BNT162b2. Compared to three weeks post-second vaccination, both cellular and humoral responses against the original SARS-CoV-2 isolate as well as variants of concern were significantly reduced, with some dialyzed individuals having no B- or T-cell response. Our data strongly support the need for a third booster in hemodialysis patients and potentially other at-risk individuals.

## Main

Persistence of vaccination-induced cellular and humoral immune responses is crucial to prevent SARS-CoV-2 infection or at least provide protection against severe COVID-19 requiring hospitalization. As in many other countries, the SARS-CoV-2 vaccination strategy in Germany was based on prioritization by occupation, underlying medical conditions or advanced age^1^. While those priority groups have been vaccinated, a debate has emerged as to whether a third “booster” dose may be necessary to maintain or raise levels of protection within some of these groups. Decisions on whether to recommend a third dose should be made within a short time frame, as it is expected that SARS-CoV-2 case numbers will increase again in the upcoming cold season, as previously observed in late 2020^2^. To date, there is however a lack of data examining the longevity of vaccination responses, with the majority of published studies only providing follow-up data until three months post-second dose^3^. Only two studies report data on extended time frames of six months after a completed two-dose scheme^4,5^ and no studies considered follow-ups in patients receiving chronic hemodialysis. Data on the actual impact of a third dose is equally scarce and so far limited to organ transplant recipients, where a third dose significantly increased antibody responses^6^. In addition, protection offered by first generation vaccines is reduced for SARS-CoV-2 variants of concern (VoC)^7^, which now represent the majority of global infections^8^ making the decision if a third dose is advisable even more critical for those with comorbidities, immunodeficiency’s or increased exposure risk such as medical staff.

One risk group for SARS-CoV-2 infection and severe COVID-19 disease are hemodialysis patients, with currently around 80,000 individuals requiring regular renal replacement therapy in Germany^9^. Their various underlying medical conditions, comorbidities, and dialysis therapy often lead to a state of generalized immunosuppression^10^. At the same time, they bear a continuous coronavirus exposure risk due to the regular need for in-centre hemodialysis therapy, which prevents them from self-isolating, or reducing contacts to avoid infection. Others and we have identified impaired cellular and humoral responses not only following SARS-CoV-2 vaccination^11-13^, but also towards other viruses such as Influenza A or Hepatitis B^10^, however there is a lack of longitudinal vaccination response studies against SARS-CoV-2 within this population. To guide future vaccination strategies if a third dose for at-risk groups for severe COVID-19 is needed, we provide follow-up data in 76 individuals receiving hemodialysis and 23 healthcare workers with no underlying conditions, for systemic and mucosal B- and T-cell responses 16 weeks after full BNT162b2 vaccination and the neutralizing potency of vaccination-induced antibodies. Due to the emergence of VoCs, which now constitute a majority of global infections^8^ and on the basis that all currently licensed vaccines are formulated against the original “wild-type (WT)” isolate (B.1), we also examined antibody binding and neutralization towards the Alpha (B.1.1.7), Beta (B.1.351), Gamma (P.3) and Delta (B.1.617.2) VoCs. As antibody levels are considered a proxy for protection, we initially examined the seroreversion rate using MULTICOV-AB^14^, a previously published bead-based multiplex immunoassay that simultaneously analyses over 20 different SARS-CoV-2 antigens including the receptor-binding domains (RBDs) of VoCs and the endemic human coronaviruses. Similarly to our previous report^11^, anti-RBD IgG responses within the dialysis group (median normalized MFI=4.26, n=76) towards SARS-CoV-2 WT RBD were significantly reduced compared to the control group (median normalized MFI=13.6, n=23, p<0.001, **Fig. 1a**) 16 weeks after complete vaccination. Compared to three weeks post-second dose (first time point), antibody titers significantly decreased by 61% in the control group and 75% in the dialysis group (p<0.001, **Fig. 1a**). Anti-RBD IgG levels measured by MULTICOV-AB were additionally verified with a commercial quantitative IVD antibody test (Spearman’s rank 0.956, **Extended Data Fig. 1a and b**). Whilst none of the samples of the control group were classified as seronegative (titer below the cut-off) (**Extended Data Fig. 2**), 19.7% (15/76) of dialysis samples were defined as such 16 weeks post-second dose, which constitutes a substantial increase from 3 weeks post-second vaccination where only 5.3% (4/76) of samples were seronegative.

**Fig. 1.**
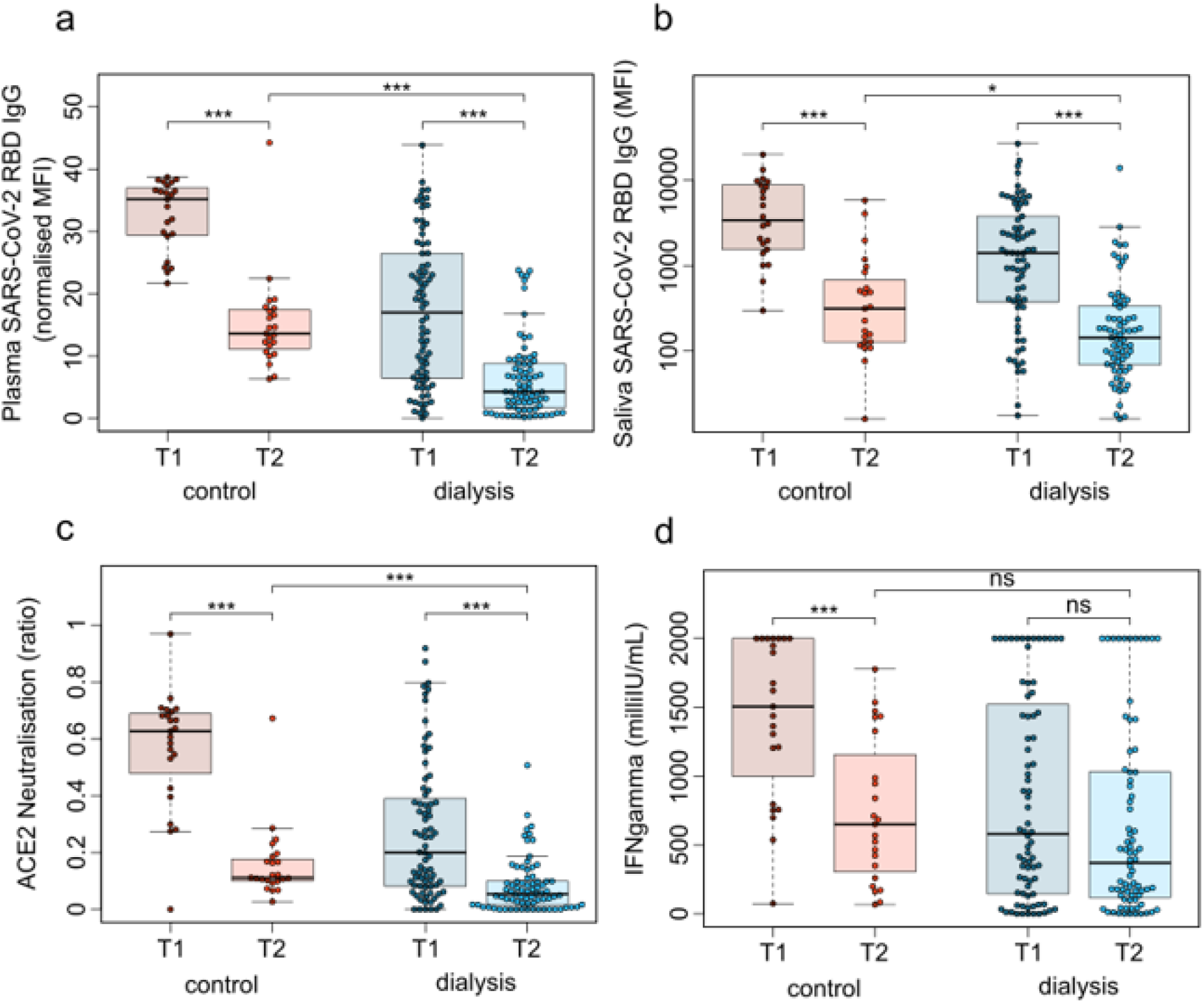
Humoral and cellular responses induced by BNT162b2 against SARS-CoV-2 significantly decrease from 3 weeks to 16 weeks post-second vaccination. IgG response in plasma (a), IgG response in saliva (b), neutralizing capacity towards SARS-CoV-2 WT B.1 (c) and T-cell response measured by IGRA (d) between dialysis (blue, n=76) and control (red, n=23) groupsSamples were taken at two time points following vaccination with Pfizer BNT162b2: T1 (three weeks post-second dose) and T2 (16 weeks post-second dose).. Saliva (b) has reduced sample numbers in both groups due to issues in sample collection (T1 control n=22, T1 dialysis n=69, T2 control n=23, T2 dialysis=71). T1 time point data has already been published^11^ and is reproduced here for clarity. Boxes represent the median, 25^th^ and 75^th^ percentiles, whiskers show the largest and smallest non-outlier values. Outliers were determined by 1.5 IQR. Statistical significance was calculated by Wilcoxon matched-pairs signed rank test when comparing between T1 and T2, and two-sided Mann-Whitney-U test when comparing between control and dialysis. Significance is defined as *** p<0.001, ** p<0.01, * p<0.05 and ns p>0.05.

To evaluate whether this reduction in plasma anti-RBD IgG was also present at the mucosal site, we profiled the local antibody response in saliva using MULTICOV-AB. As seen in plasma, there was a significant reduction in saliva anti-RBD IgG titers in the dialysis (median=143, n=71) compared to the control group (median=313.5, n=23, p=0.02, **Fig. 1b**). When comparing saliva anti-RBD IgG levels to the initial time point, there was a statistically significant decline in both groups (p<0.001, **Fig. 1b)**, suggesting that they have potentially lost competence to prevent transmission if infected. When examining anti-RBD IgA, there was a significant difference in titers between control and dialysis individuals (p=0.003, **Extended Data Fig. 3a**), with 47.8% of control and 75% of dialysis individuals classified as seronegative. This more pronounced reduction in IgA vs. IgG levels most likely represents the shorter IgA half-life. Interestingly, saliva anti-RBD IgA tended to be higher in the dialysis group although not significantly (p=0.051, **Extended Data Fig. 3b**).

We next examined whether neutralization potential was also hindered since there is solid evidence on the protective role for neutralizing serum antibodies^15^. Using an ACE2–RBD competition assay, which assesses neutralization potency towards SARS-CoV-2 wild-type (WT) and the currently circulating Alpha, Beta, Gamma and Delta VoCs, we found that neutralization against WT SARS-CoV-2 RBD was significantly reduced in the dialysis group compared to the controls (p<0.001, **Fig. 1c**) 16 weeks after complete vaccination. 82.6% (19/23) of control and 89.5% (68/76, **Extended Data Fig. 2**) of dialysis samples were below the 0.2 threshold, which indicates the absence of neutralizing activity. This 0.2 neutralization threshold is based on information provided for other available ACE2 competition assays^16^. This is a substantial significant reduction (both p<0.001) in neutralizing activity compared to 3 weeks post-second vaccination, where only 4.3% (1/23) of the control samples and 50.0% (38/76) of the dialysis samples were below the threshold (**Fig. 1c, Extended Data Fig. 2**).

As some individuals might be able to control and clear SARS-CoV-2 infections with a strong T-cell response alone, we examined Spike-specific SARS-CoV-2 T-cell responses using a commercially available Interferon ⍰ release assay (IGRA). While absolute mean IFN□ responses in the dialysis group compared to the control group tended to be lower (median 370 vs. 651 mIU/mL), this was non-significant (p=0.13, **Fig. 1d**). Interestingly, within the control group, IFN□ release after restimulation declined significantly from the first time point (median=1505, p<0.001, Fig. 1d), while for dialysis patients, this decline was non-significant (median=580, p=0.13, **Fig. 1d)**. This is likely due to the majority of control samples being at the assay’s upper limit of detection at the first time point, when the dialysis samples already showed reduced IFN□ release. Overall, the number of non-responders was higher in the hemodialysis group (40.8%, 31/76) than the control group (21.7%, 5/23, **Extended Data Fig. 2**). A lack of serological response appears to be more driven by T-cell immunity than B-cell immunity, with 2.6% (2/76) of the dialysis group having a T-cell response but no B-cell response, as opposed to 23.6% (18/76) who had a B-cell response but no T-cell response. In total, 17.1% (13/76) of the dialysis group were classified as complete non-responders due to the absence of both detectable SARS-CoV-2 wild-type B- and T-cell responses, as opposed to none in the control group.

Having characterized response against wild-type SARS-CoV-2, we finally assessed humoral response against the current VoCs (Alpha, Beta, Gamma and Delta). As shown with classical cell-culture based virus neutralization assays^7^, neutralization responses were also reduced for all VoCs when compared to WT using the previously described ACE2-RBD competition assay. Compared to the initial time point, neutralization decreased significantly for both the Alpha and Beta VoCs (both p<0.001, **Fig. 2a and b**). We were unable to determine these changes for Gamma and Delta, since these variants were not measured in the initial analysis. When comparing between the dialysis and the control cohort, dialysis individuals had significantly reduced neutralization against Alpha (p<0.001, **Fig. 2a**), Gamma (p=0.014, **Fig. 2c**) and Delta (p=0.002, **Fig. 2c**), but not for Beta (p=0.08, **Fig. 2b**). The number of non-responders was variable between the different strains although consistently high, with 87.0% of the control and 93.4% of dialysis considered non-responders against Alpha, 95.7% of control and 100% of dialysis against beta and Gamma, and 95.7% of control and 96.1% of dialysis against Delta.

**Fig. 2.**
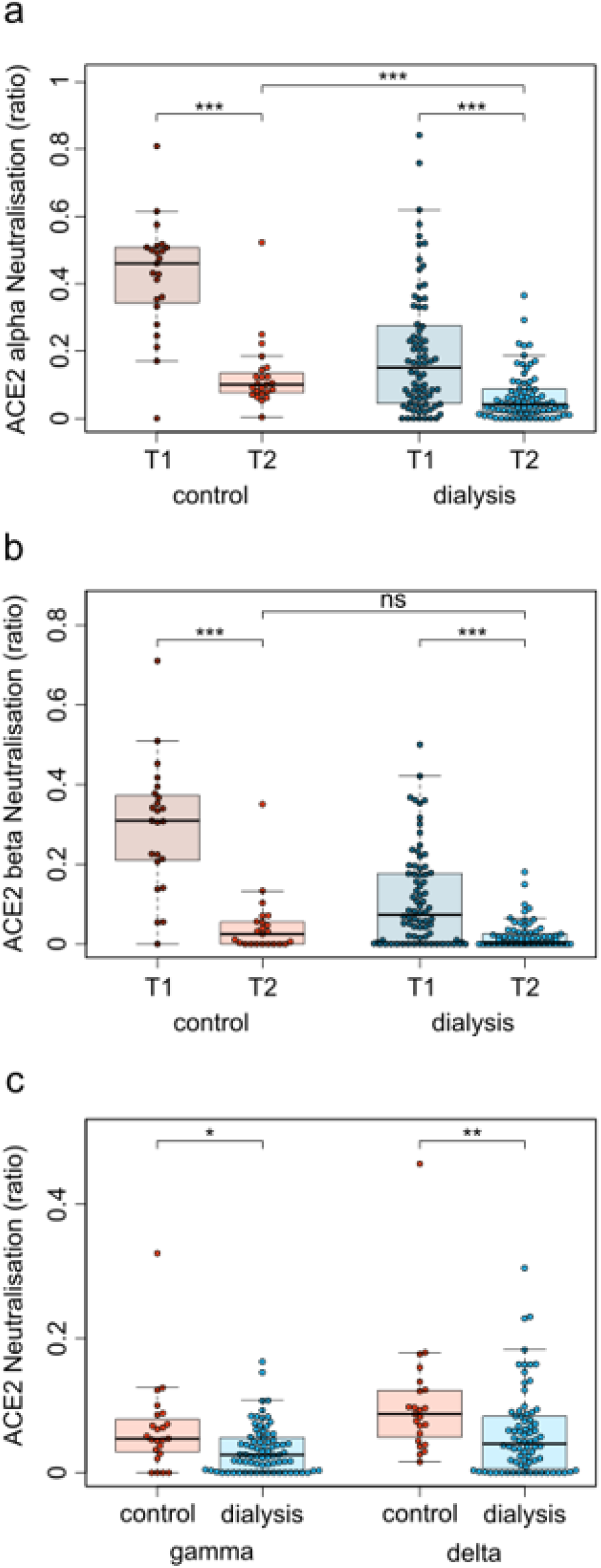
Dialysis patients have reduced neutralizing capacity against SARS-CoV-2 VOCs. Neutralizing capacity of plasma IgG towards SARS-CoV-2 VoCs Alpha (a), Beta (b), Gamma and Delta (both c) in the control (red, n=23) and dialysis groups (blue, n=76) 16 weeks post-second vaccination with Pfizer BNT162b2. Neutralization capacity is displayed as ratio where 1 indicates maximum neutralization and 0 no neutralization. Boxes represent the median, 25^th^ and 75^th^ percentiles, whiskers show the largest and smallest non-outlier values. Outliers were determined by 1.5 IQR. Statistical significance was calculated by two-sided Mann-Whitney-U test. Statistical significance is defined as *** p<0.001, ** p<0.01, * p<0.05 and ns p>0.05.

As far as we are aware, this is the first study examining the longitudinal response following vaccination, for more than 2 months post-vaccination within an immunocompromised population. In comparison to other vaccine studies, which have mostly examined peak humoral response within one month or alternative prime boost vaccination schedules with BNT162b2^13^, our data reveals a substantial decrease in the following months in hemodialysis patients and healthy controls. Overall, the decline in neutralizing anti-Spike RBD antibodies was comparable in both groups and the difference between groups mostly driven by differences in the magnitude of the initial humoral response. While this decrease is expected and can be attributed to the memory phase, the extent of the reduction was unpredicted as it resulted in a significant proportion of individuals being classified as seronegative. The reduction of salivary antibodies is particularly important as their presence has been linked to reduced transmission potential^17^. This pattern of reduced antibody binding with increasing time post vaccination was also reflected in diminishing neutralization potential. Worryingly, the majority of individuals tested were classified below our defined neutralization threshold for WT RBD with an almost complete non-responder rate against Delta, which is currently the dominant strain in many parts of the world^8^. Although this does not automatically translate to a failure of vaccine efficacy, as any active challenge of the immune system should result in expansion of memory B- and T-cell populations along with increased (neutralizing) antibody titers, it does however suggest that active protection against infection may be reduced. Whilst a recent study by Pfizer^4^ indicated that BNT162b2 vaccine efficacy did only slightly decrease six months post-vaccination in the study cohort (95% to 91%) in fully immunocompetent individuals, data from vaccinations in Israel identified a reduction in efficacy to 40%^18^. In combination with our data, where 17.1% of the dialysis cohort were classified as having no evidence for vaccine-elicited T- and B-cell immunity after four months, it is suggestive that vaccine efficacy may be even further reduced within this patient group. For dialysis patients, this is particularly concerning as they often suffer from comorbidities, which put them at additional risk for severe COVID-19^10^. The lack of a considerable SARS-CoV-2 specific T-cell response in dialysis patients may result from chronic inflammatory conditions leading to T-cell exhaustion and suppression of IFN⍰ levels^19^. Differences in anti-SARS-CoV-2 T-cell kinetics between groups presumably reflect difference in the magnitude of T-cell responses after boost and during the contraction phase. To what extend T cell immunity contributes to protection from COVID-19 and whether our IRGA results below a cut-off provide evidence for the lack of effective adaptive T-cell immunity, requires further investigation.

This study is limited by the relatively small sample size of individuals, which were not age or gender matched. However, the sample number and compromised matching is consistent with similar studies on dialysis vaccine responses^13^. While studies have indicated that differences exist in both protection and antibody responses^20^ after different COVID-19 vaccination schedules, our study of Pfizer’s BNT162b represents a real-world situation for the majority of dialysis patients. Considering reduced anti-Spike responses four weeks post-vaccination in patients with other chronic conditions^6^, these groups should undergo careful monitoring to determine whether their responses also decrease substantially over time. Taken together, our results strongly argue towards administering a third dose of BNT162b2 preferentially to all individuals undergoing chronic hemodialysis.

## Data Availability

All data is available from the corresponding authors upon request.

## Author Contributions

MS, GK, GMNB and NSM conceived the study. AD, MS, MB, DJ, NSM, A D-J, and GMNB designed the experiments. GMR, MVS, and JG performed the experiments. KL, AB, EW, GL, AC, A D-J, MVS, ADJ and GMNB collected samples and data or organized their collection. PDK, BT and UR produced and designed recombinant assay proteins. AD, MS, MB and JG performed data collection and analysis. AD and MB generated the figures. AD and MS wrote the manuscript. All authors critically reviewed and approved the final manuscript.

## Acknowledgments

We thank staff and participants at the Dialysis Centre Eickenhof for their continued support to make this study possible.

