## Supplementary material for "Diminishing immune responses against variants of concern in dialysis patients four months after SARS-CoV-2 mRNA vaccination": Online Methods

Author Affiliations

**Methods**

**Study design and sample collection**. Blood samples were collected using vascular access prior to the start of dialysis or by venipuncture for the control population 16 weeks after the standard-two dose vaccination with a 21 day interval of Pfizer BNT162b2 was completed. 76 patients on maintenance hemodialysis and 23 healthcare workers from the same dialysis center participated in the longitudinal follow-up^1^. Detailed information about the study population can be found in Extended Data Table 1, 2 and 3. Plasma was obtained from lithium heparin blood (S-Monovette, Sarstedt). Whole blood samples were used immediately for Interferon ɣ release assay. To inactivate potential pathogens, collected saliva samples were treated with Tri(n-butyl) phosphate (TnBP) and Triton X-100 for final concentrations of 0.3% and 1%, respectively.

**Ethics statement.** The study was approved by the Internal Review Board of Hannover Medical School (MHH, approval number 8973_BO-K_2020, amendment Dec. 2020). Written informed consent was obtained from all participants prior to study start.

**MULTICOV-AB.** IgG and IgA binding and levels were analyzed using MULTICOV-AB, a multiplex coronoavirus immunoassay as previously described^2^. All recombinant proteins used as MULTICOV-AB antigens in this study are listed in Extended Data Table 4. Briefly, antigens were immobilized on spectrally distinct populations of MagPlex beads (Cat #MC10XXX-01, Luminex Corporation) either by EDC/s-NHS coupling^2^ or by Anteo coupling (Cat #A-LMPAKMM-10, Anteo Tech Reagents) following the manufacturers instruction^3^. The combined MagPlex beads were then incubated with samples. After a wash step to remove unbound antibodies, IgG or IgA were detected with either R-phycoerythrin labeled goat-anti-human IgG (Jackson ImmunoResearch Labs, Cat #109-116-098, Lot #148837, RRID: AB_2337678) or IgA (Jackson ImmunoResearch Labs, Cat #109-115-011, Lot #143454, RRID: AB_2337674) as secondary antibodies. After another wash step and bead resuspension, samples were measured once on a FLEXMAP 3D instrument (Luminex Corporation) using the following settings: Timeout 80 sec, Gate: 7500-15000, Reporter Gain: Standard PMT, 40 events. Raw median fluorescence intensity (MFI) values or normalized values (MFI/MFI of quality control (QC) samples^3^ are reported. Three QC samples were measured per individual plate to monitor MULTICOV-AB performance.

**ACE2-RBD competition assay**. An ACE2-RBD competition assay was carried out as previously described^3^ to determine immunoglobulin neutralization potency. For this, biotinylated ACE2 was combined with individual samples (and as a control, ACE2 alone) and incubated with the above mentioned MULTICOV-AB bead mix. Before and after ACE2 detection with Strep-PE (Cat #SAPE-001, Moss), washes were carried out. Samples were measured once on a FLEXMAP 3D instrument with the same settings as MULTICOV-AB and analyzed by normalization of MFI values against the control.

**Interferon γ release assay (IGRA).** SARS-CoV-2-specific T-cell responses from whole blood were analyzed by measuring IFNγ production after stimulation with a peptide pool from the SARS-CoV-2 Spike S1 with the SARS-CoV-2 Interferon Gamma Release Assay (Cat #ET-2606-3003, Euroimmun) and the IFNɣ ELISA (Cat #EQ-6841-9601, Euroimmun) as previously described^1^. Background signals from negative controls were subtracted and final results calculated in mIU/mL using standard curves. Results from positive and negative controls were not statistically significant different between time point T1 and T2. IFNγ concentrations >200 mIU/mL were considered as reactive. We defined this arbitrary cut-off by using average background IFNγ activity without antigen-stimulation in all samples of T1 multiplied with 10 for the threshold for IGRA-positive. Using this cut-off, we found in all of the 15 controls taken from independent individuals before the COVID-19 pandemic negative IGRA results^4^. The upper limit of reactivity was 2000 mIU/mL.

**QuantiVac-SARS-CoV-2 ELISA.** Plasma samples were additionally analyzed using the Anti-SARS-CoV-2-QuantiVac-ELISA IgG (Cat# EI 2606-9601-10G, Euroimmun) as previously described^1^.

**Statistical analysis**. Matching sample metadata and collecting results from different assay platforms was performed in Excel 2016. GraphPad Prism 8.4.3 was used for statistical analysis. Figures were generated in RStudio (Version 1.2.5001) running R (version 3.6.1). The add-on package “beeswarm” was utilized to visualize data as strip charts with overlaying boxplots and to create non-overlaying data points. “RcolorBrewer” add-on was used to generate specific colors for plots. Figures were then edited Inkscape (Inkscape 0.92.4). The type of statistical analysis used is listed in the corresponding figure legend.

**Data availability**

Data is available from the corresponding authors upon request.
