## Extended Data for "Diminishing immune responses against variants of concern in dialysis patients four months after SARS-CoV-2 mRNA vaccination"

Author Affiliations

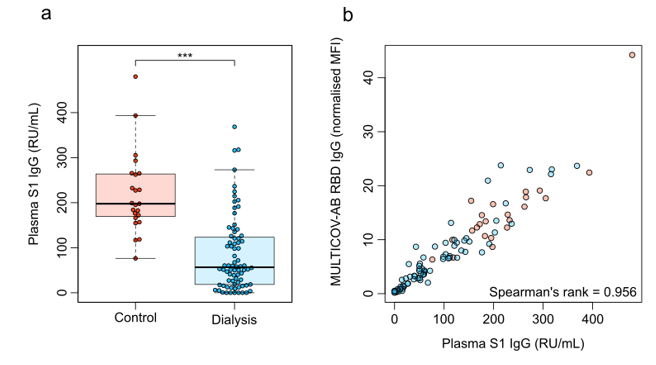

**Extended Data Fig. 1 | Quantitative plasma IgG titers 16 weeks after vaccination with Pfizer BNT162b2.**

(a) Spike S1-specific plasma IgG (RU/mL) from control group (red, n=23) and dialysis group (blue, n=76) were analyzed 16 weeks post-second dose of Pfizer BNT162b2 using the QuantiVac-ELISA (Euroimmun). Samples above upper or below the ELISA’s limits of detection are shown at the corresponding limit. Boxes represent the median, 25^th^ and 75^th^ percentiles, whiskers show the largest and smallest non-outlier values. Outliers were determined by 1.5 times IQR. Statistical significance was calculated by two-sided Mann-Whitney-U test. Significance was defined as ***<0.001. (b) Correlation of MULTICOV-AB wild-type RBD B.1-IgG and QuantiVac Spike S1-IgG across the study population. Spearman’s rank was used for correlation analysis.

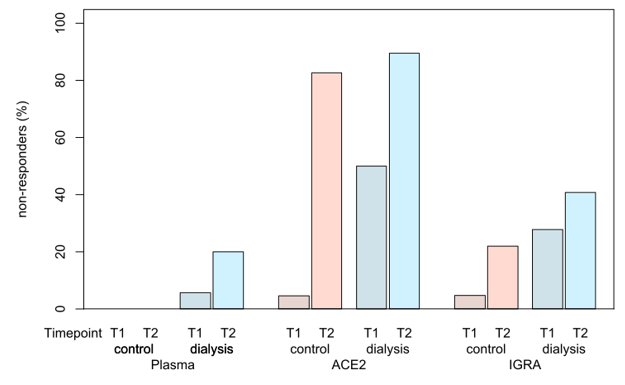

**Extended Data Fig. 2 | B- and T-cell non-responder rate at 3 weeks post-vaccination (T1) and 16 weeks post-vaccination (T2).**

Non-responders (displayed as % of the total) for plasma IgG (MULTICOV-AB), ACE2 neutralization (ACE2-RBD competition assay) and IFNɣ release (IGRA). Data is shown as boxplots. T1 data has already been published and is reproduced here for clarity^1^.

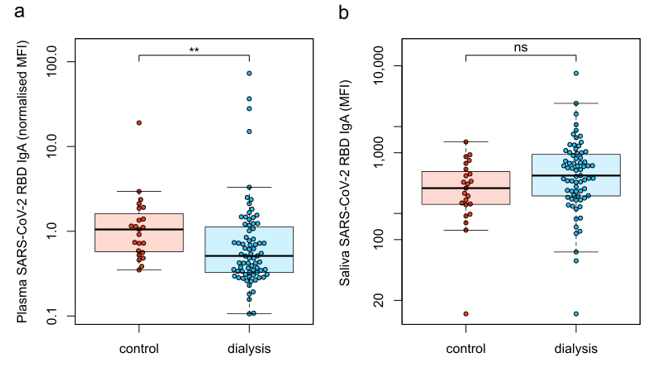

**Extended Data Fig. 3 | IgA response 16 weeks after vaccination with Pfizer BNT162b2.**

IgA binding in plasma (a) and saliva (b) towards the RBD of SARS-CoV-2 WT is displayed as MFI using MULTICOV-AB. Samples of control group (red, n=23) and dialysis group (blue, n=76) were collected 16 weeks after the second dose of Pfizer BNT162b2. Sample numbers for saliva (b) in the dialysis group are reduced to 72. Boxes represent the median, 25^th^ and 75^th^ percentiles, whiskers show the largest and smallest non-outlier values. Outliers were determined by 1.5 times IQR. Statistical significance was calculated by two-sided Mann-Whitney-U test. Significance was defined as *<0.05, **<0.01, ***<0.001 or n.s.>0.05.

**Extended Table 1.** Characteristics of vaccinated study participants.

|  | Non-dialysis control group (n=23) | Hemodialysis group (n=76) | p-value for difference between groups |
| --- | --- | --- | --- |
| Age (years), median (IQR) | 55 (14) | 70.5 (18.25) | 2.78 x 10^-9^ |
| Gender (female, n, %) | 17 (73.91) | 33 (43.42) | 1.01 x 10^-2^ |
| Days since start of hemodialysis (median, IQR) | n. a. | 1337 (1686.5) | n. a. |
| Immunosuppressive medication (n, %) | 0 (0) | 10 (13.16) | 6.77 x 10^-2^ |
| Co-morbidities |  |  |  |
| Obesity (BMI, >30.0) | 4 (17.39), 1 NA | 16 (21.05) | 8.68 x 10^-1^ |
| Diabetes mellitus (n, %) | 0 (0) | 19 (25) | 7.30 x 10^-3^ |
| Cardiovascular disease (n, %) | 0 (0) | 35 (46.05) | 2.93 x 10^-5^ |

IQR: Inter Quartile Range. BMI: Body Mass Index. n – absolute numbers per group. NA: Information not available. n. a.: not applicable.

**Extended Table 2.** Therapeutic indication for hemodialysis.

| Characteristics | Hemodialysis group (n=76) |
| --- | --- |
| Diagnosis (n, %) |  |
| Autosomal dominant polycystic kidney disease | 11 (14.47) |
| Chronic glomerulonephritis | 6 (7.90) |
| Diabetic nephropathy | 11 (14.47) |
| Focal segmental glomerulosclerosis | 5 (6.59) |
| IgA nephropathy | 8 (10.53) |
| Interstitial nephropathy | 6 (7.90) |
| Nephrosclerosis | 16 (21.05) |
| Acute toxic tubular epithelial damage syndrome | 1 (1.32) |
| Primary amyloidosis | 1 (1.32) |
| [ANCA-associated vasculitis](https://www.bmj.com/content/369/bmj.m1070) | 1 (1.32) |
| Cardiorenal syndrome | 2 (2.64) |
| Medullary cystic kidney disease | 1 (1.32) |
| Membranous glomerulonephritis | 1 (1.32) |
| Kidney dysplasia | 1 (1.32) |
| Obstructive nephropathy | 1 (1.32) |
| Reflux nephropathy | 1 (1.32) |
| Septic organ failure | 1 (1.32) |
| Cystic kidney disease | 1 (1.32) |
| Cyclosporin intoxication | 1 (1.32) |

ANCA: Anti-Neutrophilic Cytoplasmic Autoantibody.

**Extended Table 3.** Medication of vaccination cohort.

| Characteristics | Non-dialysis control group (n=23) | Hemodialysis group (n=76) |
| --- | --- | --- |
| Medication (n, %) |  |  |
| Angiotensin-converting enzyme inhibitors | 2 (8.70) | 22 (28.95) |
| Statins | 0 (0) | 45 (59.21) |
| Angiotensin II Receptor Blockers | 5 (21.74) | 25 (32.89) |
| Vitamin D Supplements | 12 (52.17) | 75 (98.68) |
| Immunosuppressants (dosing range per day)** |  |  |
| Prednisolone (2-7.5 mg) | 0 | 5 (6.59) |
| Prednisolone (50 mg) day 6-14 post 2nd vaccination | 0 | 1 (1.32) |
| Prednisolone (5 mg), Tacrolimus (0·5-2 mg) | 0 | 2 (2.64) |
| Prednisolone (5 mg), Tacrolimus (12 mg), Mycophenolatmofetil (500 mg) | 0 | 1 (1.32) |
| Hydrocortisone (20 mg) | 0 | 1 (1.32) |

**Therapeutic indication for immunosuppression were in four patients a kidney transplant (one had received an additional liver transplant), polymyositis, polyarthritis, vasculitis and chronic obstructive pulmonary disease.

**Extended Table 4.** MULTICOV-AB antigen panel.

| Virus | Antigen | Manufacturer | Product number |
| --- | --- | --- | --- |
| SARS-CoV-2 | Spike Trimer | NMI | - |
| SARS-CoV-2 | RBD B.1 (wild-type) | NMI | - |
| SARS-CoV-2 | Nucleocapsid | Aalto | 6404-b |
| SARS-CoV-2 | RBD B.1.1.7 (Alpha) | NMI |  |
| SARS-CoV-2 | RBD B.1.351 (Beta) | NMI |  |
| SARS-CoV-2 | RBD P.3 (Gamma) | NMI |  |
| SARS-CoV-2 | RBD B.1.617.2 (Delta) | NMI |  |
| hCoV-OC43 | S1 domain | NMI | - |
| hCoV-OC43 | Nucleocapsid | NMI | - |
| hCoV-HKU1 | S1 domain | NMI | - |
| hCoV-HKU1 | Nucleocapsid | NMI | - |
| hCoV-NL63 | S1 domain | NMI | - |
| hCoV-NL63 | Nucleocapsid | NMI | - |
| hCoV-229E | S1 domain | NMI | - |
| hCoV-229E | Nucleocapsid | NMI | - |
